# Delayed humoral kinetics but stabilization of IgG responses in common variable immunodeficiency after SARS-CoV-2 mRNA booster vaccination

**DOI:** 10.64898/2026.05.07.26352649

**Authors:** Lorenzo Federico, Ragnhild Øye Løken, Quy Khang Le, Julie Røkke Osen, Viktoriia Chaban, Ingvild Nordøy, Tonje Skarpengland, Knut Erik Aslaksen Lundin, Silje Fjellgård Jørgensen, Mai Sasaki Aanensen Fraz, Pål Aukrust, Katrine Persgård Lund, Trung The Tran, Liv Toril Nygård Osnes, Fridtjof Lund-Johansen, Hassen Kared, Børre Fevang, Ludvig Andre Munthe

## Abstract

**Purpose:** Patients with common variable immunodeficiency (CVID) frequently exhibit impaired antibody responses to vaccination, yet the dynamics of humoral and cellular immunity following mRNA immunisation remain incompletely defined. This study aimed to characterise the temporal evolution of anti-SARS-CoV-2 antibody and T cell responses across successive vaccine doses in a well-characterised CVID cohort, and to identify key determinants of vaccine responsiveness in this population.

**Methods:** We performed a longitudinal and cross-sectional analysis of serum and peripheral blood mononuclear cell (PBMC) samples collected from 88 CVID patients after two, three, or four doses of mRNA vaccine (Moderna/mRNA-1273 or Pfizer-BioNTech/BNT162b2). Anti-receptor-binding domain (RBD) IgG titers were quantified in relation to vaccine dose, time since last vaccination, and clinical characteristics. Vaccine-specific CD4^+^ and CD8^+^ T cell responses were assessed *ex vivo* using an activation-induced marker (AIM) assay by flow cytometry.

**Results:** The proportion of patients with detectable anti-RBD IgG increased from 35% after two doses to more than 80% after four doses. Boosting-dependent increases in IgG titers were observed exclusively in samples collected more than three months after the last dose, and antibody levels correlated positively with time since vaccination, consistent with delayed but progressive humoral kinetics that stabilised after the third dose. In contrast, spike-specific CD4^+^ and CD8^+^ T cell responses were rapidly induced and remained stable across all timepoints.

**Conclusion:** Vaccine-induced immunity in CVID is characterised by delayed humoral responses alongside preserved cellular immunity. Early post-vaccination serology may systematically underestimate vaccine responsiveness, and booster vaccination supports stabilisation of antibody responses in this population.

## Introduction

Common variable immunodeficiency (CVID) is the most prevalent symptomatic primary antibody immunodeficiency in adults and is characterized by impaired immunoglobulin production and defective B-cell differentiation, leading to recurrent respiratory infections with encapsulated bacteria, increased susceptibility to certain viral infections, and predisposition to severe outcomes following SARS-CoV-2 infection [1]. In addition, 70–80% of CVID patients develop autoimmune, inflammatory, or lymphoproliferative complications, which are associated with increased morbidity and mortality compared to patients presenting with infections alone [2, 3].

Although the underlying immunopathology is heterogeneous [4, 5], patients exhibit impaired class-switch recombination, with a notable fraction also showing defective germinal center formation [6, 7], contributing to their reduced capacity to mount protective immune responses to infections [8] and vaccination [9–11]. However, abnormalities in T cells and monocytes are frequently observed, especially in patients with autoimmune, inflammatory, and lymphoproliferative complications, and T cell dysfunction may contribute to B cell defects via impaired T–B cell interactions [12, 13].

mRNA-based SARS-CoV-2 vaccines elicit robust immune responses in healthy individuals but more variable responses in immunocompromised populations [14–17]. Several studies have reported that following vaccination, CVID patients exhibit diminished seroconversion rates with attenuated antibody titers [18–20], but generally sustained T cell responses that are thought to contribute significantly to protection from severe infection [21–23]. However, most studies report assessment of immune responses at diverse time points and doses of vaccine [23–26]. The heterogeneity of these observations—driven for the most part by variable intervals between vaccination and sample collection—may confound the interpretation of vaccine-response kinetics and limit our understanding of immune-response dynamics in CVID. Consequently, additional data on the time-dependent evolution of serological responses and the interplay between specific immune-cell subsets and their activation status are warranted to better inform clinical assessment of vaccine responsiveness and to optimize vaccination strategies.

We hypothesized that patients with CVID would exhibit not only an attenuated immune response to the SARS-CoV-2 vaccine but also altered response kinetics. To better define these immune-response dynamics, we performed an integrated analysis of humoral and cellular immunity following SARS-CoV-2 mRNA vaccination in a cohort of CVID patients whose serum and peripheral blood mononuclear cells (PBMCs) were collected after two or more vaccine doses administered during the pandemic and subsequent endemic period. Anti–receptor-binding domain (RBD) IgG titers were quantified according to vaccine dose, sampling interval post-vaccination, clinical characteristics, and peripheral B and T cell subset composition, while vaccine-specific CD4^+^ and CD8^+^ T cell responses were assessed *ex vivo* by flow cytometry using an activation-induced marker (AIM) assay.

Consistent with previous observations, CVID patients exhibited reduced antibody responses after vaccination; however, both boosting and time since vaccination were key determinants of detectable humoral immunity. Despite impaired humoral responses, spike-specific CD4^+^ and CD8^+^ T cell responses were rapidly induced and remained stable over time. Together, these findings indicate that booster vaccination stabilizes humoral immunity in CVID and that preserved cellular immunity may contribute to protection when humoral responses are not fully established.

## Methods

### Study design and participants

Peripheral blood samples were obtained after SARS-CoV-2 vaccination from 88 individuals diagnosed with CVID according to European Society for Immunodeficiencies (ESID) 2019 criteria who had received at least two doses of mRNA vaccines (Moderna/mRNA-1273 or Pfizer-BioNTech/BNT162b2) between 21 May 2021 and 1 December 2022. CVID patients were classified as “Infection only” or “Autoimmune and inflammatory complications” (“Complications”) phenotype according to Chapel et al. [27]. Clinical characteristics and sample availability are summarized in Table 1. In addition, samples from six patients with X-linked agammaglobulinemia (XLA) and five individuals with primary unspecific hypogammaglobulinemia (puHG) were included. All patients received immunoglobulin replacement therapy, administered intravenously (IVIG), subcutaneously (SCIG), or via a combination of both routes. The interval between IVIG infusion and serum sampling ranged from 0 to 4 weeks. For SCIG-treated patients, sampling times were not recorded because serum anti-RBD IgG concentrations are relatively stable when immunoglobulins are administered via this route. Serum samples from healthy donors (n = 333) [16] were used as controls. Vaccine-induced T cell responses were evaluated *ex vivo* in cryopreserved PBMCs from CVID patients (n = 29 pre-vaccination; n = 16 after two doses; n = 7 after three doses) and compared with PBMCs from a previously established biobank of healthy individuals (n = 57) who had received two or three doses of Moderna (mRNA-1273) or Pfizer–BioNTech (BNT162b2). The study protocol and all associated biobanking procedures conducted at Oslo University Hospital were approved by the Norwegian Regional Committee for Medical and Health Research Ethics (approval number 233704) and by the Data Protection Officer of the Oslo University Hospital. Written informed consent was obtained from all participants.

**Table 1.**
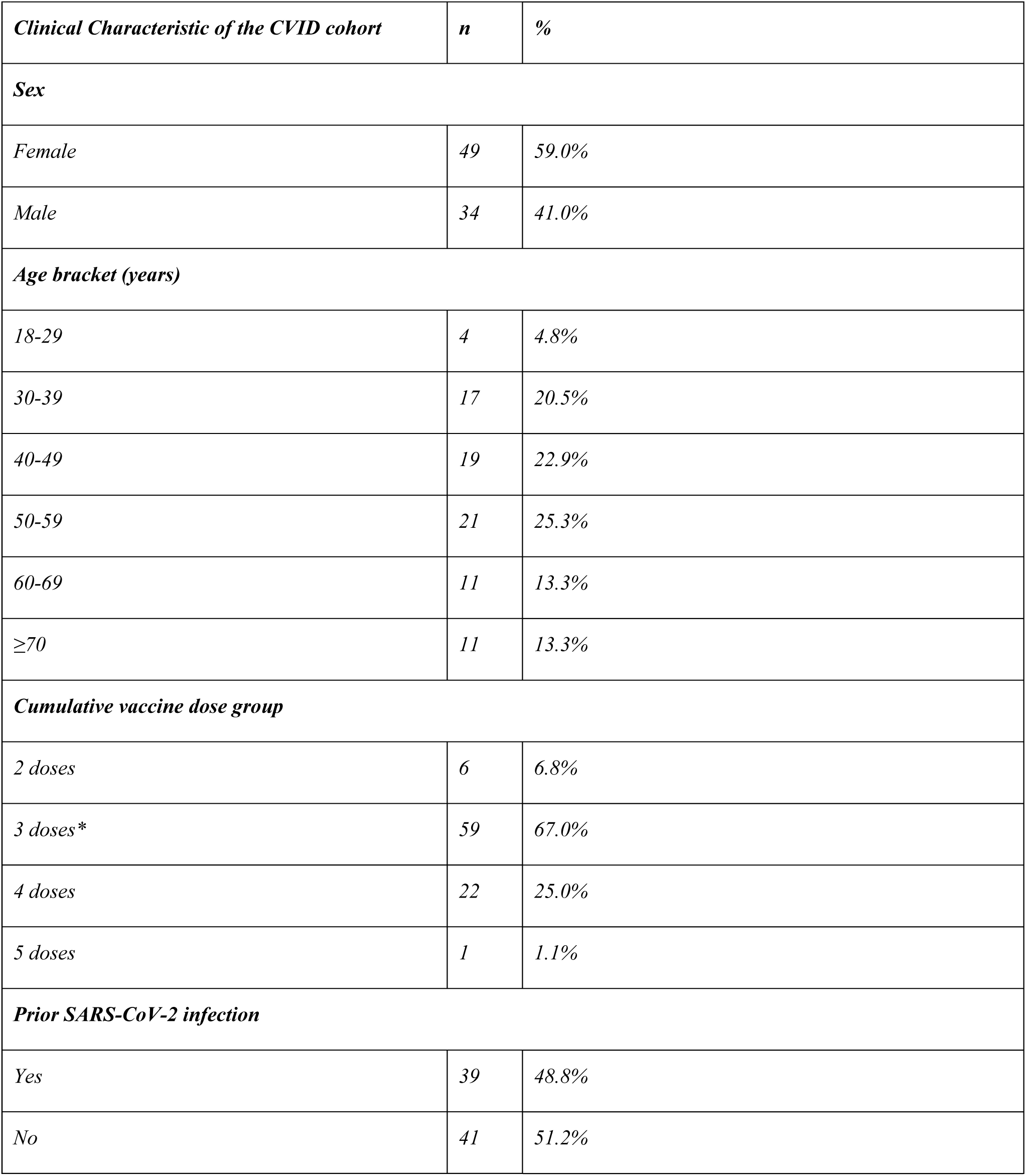
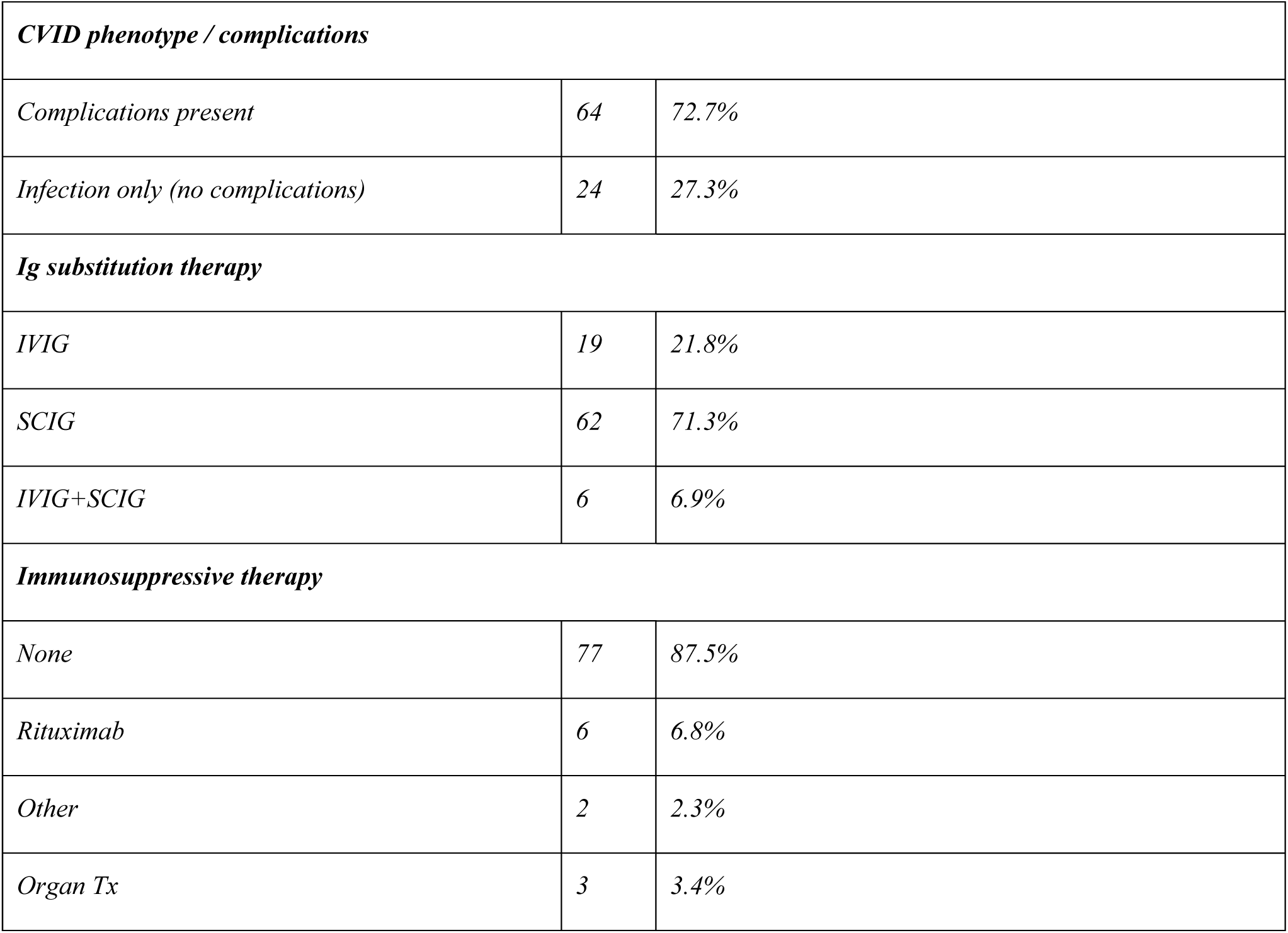
Clinical characteristics of the CVID cohort. Summary of clinical characteristics and serum anti-RBD IgG measurements for all samples included in the study. Percentages exclude missing values. A total of 88 unique patients were included (≥1 valid anti-RBD IgG measurement, BAU/mL). Cumulative dose groups indicate the maximum number of vaccine doses received per patient. *The subgroup (n = 59) includes 44 patients sampled only after the third vaccine dose, as they were enrolled after receiving two doses. Abbreviations: CVID, common variable immunodeficiency; IVIG, intravenous immunoglobulin; SCIG, subcutaneous immunoglobulin.

### Serum and PBMC isolation

PBMCs were isolated from whole blood using either BD Vacutainer® CPT tubes (#362782) or density gradient centrifugation with Lymphoprep™ (#1858). CPT tubes were centrifuged at 1,600 × g for 25 min at room temperature, after which serum was collected and stored at −20°C for subsequent analyses. Lymphoprep™ separation was performed according to the manufacturer’s protocol. Briefly, whole blood was diluted 1:1 with phosphate-buffered saline (PBS), layered onto 10–15 mL of Lymphoprep™ in 50-mL conical tubes, and centrifuged at 800 × g for 25 min. PBMCs were harvested, washed in PBS (Gibco, #10010-015), and centrifuged three additional times (400 × g, 7 min, 4°C). Cells were counted and resuspended in fetal bovine serum (FBS; Gibco, #10270-106) supplemented with 10% dimethyl sulfoxide (DMSO). Following overnight cooling at −80°C in a controlled-rate freezing container (Mr. Frosty™, Nalgene™, #5100-0001), samples were transferred to liquid nitrogen for long-term storage.

### Immunophenotyping

Routine enumeration of B and T cell lymphocyte subsets was performed on EDTA-anticoagulated peripheral blood samples by flow cytometry at the Department of Immunology, Oslo University Hospital Rikshospitalet. Absolute counts of B cells, T cells, and NK cells were determined using Trucount™ tubes (BD Biosciences, Franklin Lakes, NJ, USA) on a FACSCanto II flow cytometer and analyzed with BD FACSCanto™ Clinical Software according to the manufacturer’s instructions. Instrument settings were standardized as recommended, and daily quality control was performed using CS&T beads and 7-color Setup Beads (BD Biosciences).

Relative frequencies of B and T cell subpopulations were assessed using a Gallios flow cytometer (Beckman Coulter, San Diego, CA, USA). Briefly, washed samples were used for B cell analyses, whereas unwashed samples were used for T cell analyses. Samples were incubated with optimally titrated fluorochrome-conjugated antibodies for 15 minutes at room temperature, followed by red blood cell lysis using BD FACSLysing Solution. Data were acquired and analyzed using Kaluza software (Beckman Coulter). For T cell analyses, 1 × 10^5^ events were acquired, while up to 1 × 10^6^ events were acquired for B cell analyses when feasible. Normal reference ranges were established using samples from healthy blood donors. The laboratory operates according to standard operating procedures and is ISO certified. T and B cell subpopulations were defined by the following markers: CD3^+^, CD4^+^, CD127−, CD25^+hi^ (Tregs); CD3^+^, CD4^+^, CD45RO^+^, CXCR5^+^ (Tfh); CD19^+^, IgM−, CD27^+^ (class-switched memory B cells); CD19^+^, CD38^++−^, IgM^++^ (transitional B cells); and CD19^+^, CD38^+lo^, CD21−/+lo (CD21low B cells).

### Serological analysis

Serum IgG antibodies directed against the SARS-CoV-2 receptor-binding domain (RBD) and nucleocapsid protein were quantified using a multiplex bead-based assay, as previously described [16]. Whole blood was collected in 5-mL serum separator tubes (Vacuette®, #456073R), centrifuged at 2,000 × g, and serum aliquots were stored at −20°C. Semi-quantitative assessment of IgG antibodies against full-length spike protein (Spike-FL) and RBD was performed using fluorescently barcoded polymer beads sequentially coupled to neutravidin (Thermo Fisher) and biotinylated viral antigens. Serum samples were diluted 1:100 in assay buffer (PBS containing 1% Tween-20, 10 µg/mL D-biotin, 10 µg/mL neutravidin, and 0.1% sodium azide) and incubated with the bead arrays in 384-well plates for 30 min at 22°C with continuous agitation. After washing with PBS-Tween (PBT), bound IgG was detected using R-phycoerythrin (R-PE)–conjugated goat anti-human IgG (Jackson ImmunoResearch). Neutralizing antibody activity was assessed by subsequent incubation with digoxigenin-labeled human ACE2 followed by R-PE–conjugated anti-digoxigenin monoclonal antibody. Beads were acquired on an Attune NxT flow cytometer (Thermo Fisher), and data were analyzed using WinList 3D software. Median fluorescence intensity (MFI) values were normalized to neutravidin-only control beads to generate relative MFI (rMFI). Seropositivity thresholds were established using 979 pre-pandemic sera and 810 convalescent COVID-19 samples. A dual cutoff of rMFI >5 for anti-RBD and anti-spike-FL provided 99.7% specificity and 95% sensitivity. Antibody concentrations were expressed in binding antibody units per milliliter (BAU/mL) using serum from a triple-vaccinated individual as a reference standard.

### SARS-CoV-2 peptide pools used in the study

T cell stimulation was performed using two 15-mer peptide libraries (11-amino-acid overlap). One pool (PepTivator® SARS-CoV-2 Prot_S; Miltenyi, #130-126-700) encompassed immunodominant regions of the spike protein, while the second pool (PepTivator® SARS-CoV-2 Prot_S Complete; Miltenyi, #130-127-953) spanned the full spike sequence (amino acids 5–1273; GenBank MN908947.3). The combined preparation was referred to as the “PepTivator mix” and used for all stimulation assays.

### T cell Activation-Induced Marker (AIM) Assay and flow cytometry

Antigen-specific T cell activation was assessed using an AIM assay adapted from published protocols. PBMCs were washed in cold RPMI-1640 supplemented with GlutaMAX™ and enriched for viable cells using magnetic separation columns (MACS MultiStand with OctoMACS™ Separator). Cells were resuspended at 10 × 10^6^ cells/mL and plated in 96-well round-bottom plates (200 µL per well) in TexMACS medium supplemented with sodium pyruvate, non-essential amino acids, 1-thioglycerol, gentamycin, and recombinant IL-2. Following a 3-h pre-incubation, cells were washed and stimulated for 2 h with the PepTivator mix in the presence of anti-CD28/CD49d co-stimulatory antibodies. Cells were then incubated for an additional 18 h with brefeldin A and monensin (GolgiStop™). After stimulation, cells were stained with a fixable Near-IR viability dye, fixed and permeabilized using BD Cytofix/Cytoperm reagents, and labelled with fluorochrome-conjugated antibodies against surface and intracellular markers, including CD3, CD4, CD8, CD137, CD40L, IFN-γ, TNF, IL-2, granzyme B, and perforin. Samples were acquired on an Attune NxT flow cytometer, and data were analyzed using FlowJo software.

### Polyfunctionality analysis

Antigen-responsive T cells were identified using Boolean gating strategies applied to combinations of activation and effector markers. Responses were defined as cells positive for two or more markers including CD137, CD154, IFN-γ^+^, IL-2^+^, and TNF^+^. Frequencies of antigen-specific cells and fractional enrichment in polyfunctional cells were calculated after subtraction of unstimulated background controls.

### Statistical analysis

Statistical analyses were performed using Python (scipy v1.17) and GraphPad Prism (v10.4.1). Normality was assessed using the Shapiro–Wilk test prior to selecting statistical methods. Non-parametric tests (Mann–Whitney U, Wilcoxon matched-pairs signed-rank, and Kruskal–Wallis) or parametric equivalents (ANOVA) with Dunn’s or Dunnett’s multiple comparisons test were applied as appropriate. Correlations were assessed using Pearson’s r for normally distributed variables, Spearman’s ρ for non-normal data, or Kendall’s τ-b where non-normality was combined with a high proportion of tied values. Multiple comparisons between anti-RBD IgG serum levels and immunophenotyping parameters were corrected using the Benjamini–Hochberg false discovery rate (FDR) method across 11 immunophenotyping parameters within each subgroup; significance threshold: FDR-adjusted p < 0.05.

## Results

### Slow but boostable response to SARS-CoV-2 mRNA vaccination in CVID patients

To evaluate SARS-CoV-2 vaccine–induced immune responses in CVID, serum anti-RBD IgG levels were quantified in patients who had received two, three, or four mRNA vaccine doses. Clinical characteristics are summarized in Table 1. Longitudinal analysis of anti-RBD IgG levels revealed marked heterogeneity in vaccine responses among CVID patients. As expected, stratification by vaccine dose number demonstrated a stepwise increase in antibody levels with successive booster doses (Supplementary Figure S1). In the subset of four patients with longitudinal samples available after the second dose, the estimated IgG half-life was 68 days (±5.5 days, SD). Given that complete longitudinal data were unavailable for the full cohort, we performed a cross-sectional analysis stratified by vaccine dose (Figure 1). When the peak antibody titer recorded for each patient was plotted regardless of sampling timepoint, only 35% of the 26 CVID patients showed detectable serum anti-RBD IgG levels (>200 BAU/mL) following two doses (V2), confirming a broadly impaired humoral response in this cohort (median 31 BAU/mL, range 0.5–6,463 BAU/mL; Figure 1A). Additional booster doses resulted in a progressive increase in the proportion of seropositive individuals, with 57% and 83% of patients achieving detectable response after the third and fourth doses, respectively (1576 BAU/mL median, 0.5–18,675 BAU/mL range after V4, Figure 1A). CVID patients (n = 86) exhibited significantly lower anti-RBD IgG titers compared to healthy (n = 333) controls after 2 or 3 doses of vaccine (50% vs. 100% seropositive, with 242.5 vs. 7103 BAU/mL median antibody levels, P < 0.0001; Figure 1B); 23 of 86 patients were still classified as seroconverted strong responders (>2000 BAU/mL). Moreover, following three vaccine doses and breakthrough infection (BTI), anti-RBD IgG titers increased significantly (761.9 vs 6543 BAU/mL; P < 0.0001, Figure 1C), with nearly 90% of patients achieving a detectable humoral response. To better characterize the dynamics of the humoral response, we stratified patients by sampling time (days since the most recent dose) dichotomized at 90 days (<3 months, “early”; >3 months, “late”) and found that significant differences in anti-RBD IgG titers following sequential vaccination were detectable only in samples collected >3 months after vaccination (Figure 1D–E). In this late-collection group, increases in anti-RBD IgG titers were observed following sequential boosting when comparing levels after the second versus the third and fourth doses (V2 vs V3, P = 0.0014; V2 vs V4, P = 0.0002, Figure 1E). Anti-RBD IgG levels correlated with time since the last dose (Spearman’s r = 0.63, P < 0.0001; third dose; Figure 1F, left), and this correlation remained significant after excluding individuals with prior SARS-CoV-2 infection (Spearman’s r = 0.56, P = 0.0034; Figure 1F, right). These results indicate that, although responses were impaired after two vaccine doses, the third dose markedly increased the proportion of responders and stabilized anti-RBD IgG titers, allowing antibodies to remain detectable at later time points.

**Figure 1.**
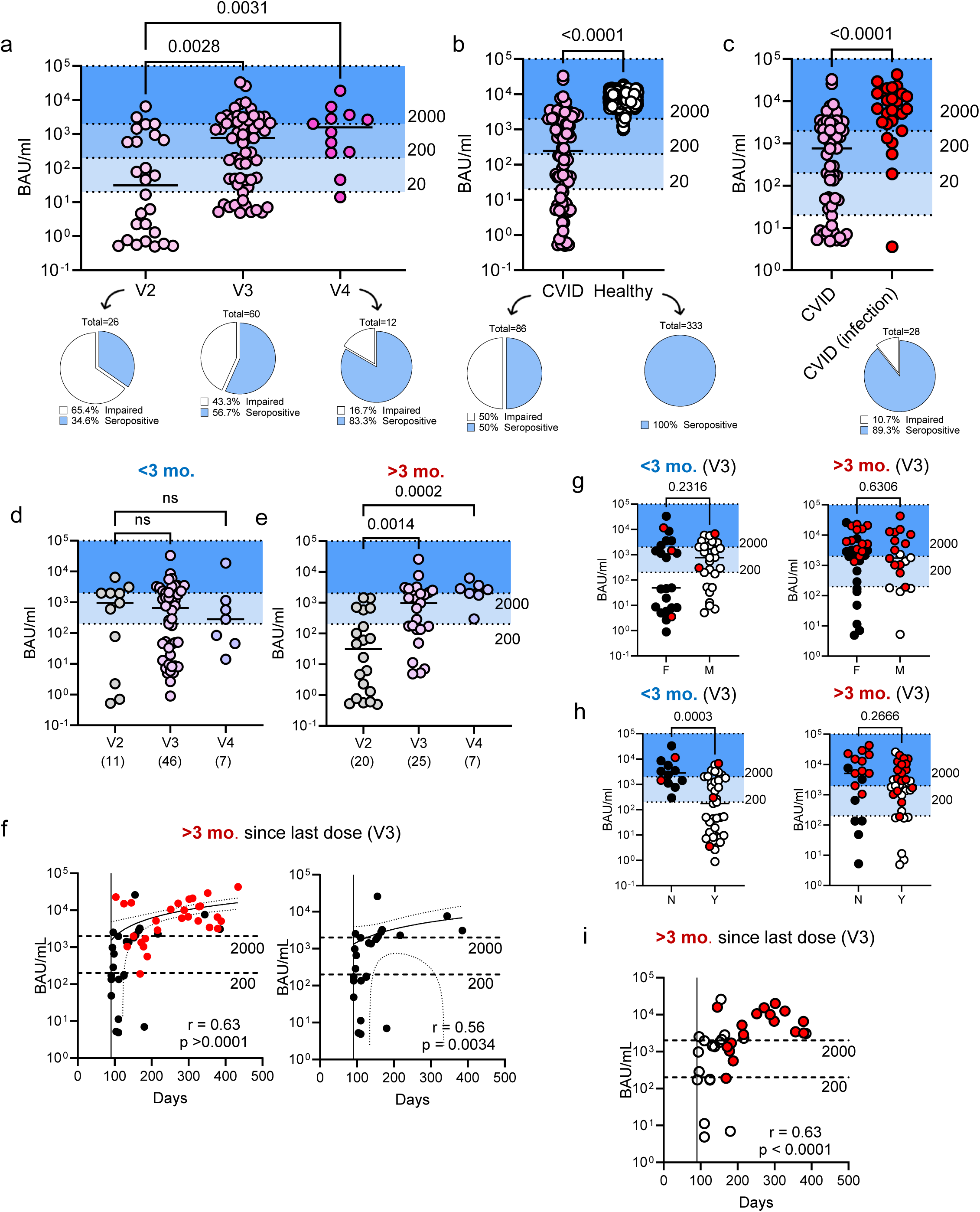
Slow but boostable response to SARS-CoV-2 mRNA vaccination in CVID patients. (A) Humoral response in CVID patients after 2, 3, and 4 vaccine doses (V2–V4). (B) Anti-RBD IgG (BAU/mL) levels in CVID patients (without prior infection) and healthy donors. All participants received either two or three doses. The median is indicated. Blue-shaded areas denote seropositive individuals (>200 BAU/mL) and strong responders (>2000 BAU/mL). (C) Comparison between CVID patients without and with prior infection after three doses of vaccine. (D–E) Seroconversion in CVID patients after 2, 3, and 4 doses, stratified by serum collection time: earlier (D) or later (E) than 3 months post-vaccination. (F) Correlation between anti-RBD IgG levels and time since the last vaccine dose (V3) in patients sampled >3 months post-vaccination. Correlation is shown including (left) and excluding (right) individuals with prior SARS-CoV-2 infection. Spearman’s r and P values are shown. (G) Sex differences in anti-RBD IgG (BAU/mL) levels. (H) Anti-RBD IgG in CVID patients with (Y) or without (N) complications. Panels show samples collected before (left panel) or after (right panel) 3 months from the third vaccination (V3). (I) Correlation between anti-RBD IgG levels and time since the last vaccine dose in patients with complications sampled >3 months post-vaccination. Statistical comparisons were performed using the Kruskal–Wallis test for panels (A), (D), and (E), and the Mann–Whitney test for panels (B), (C), (G), and (H). ns, not significant. A 95% confidence band is shown in panels (F) and (I). The number of patients is indicated in parentheses. Red dots represent anti-RBD IgG levels measured after reported SARS-CoV-2 infection. BAU, binding antibody units.

Sex did not impact seroconversion (Figure 1G), but CVID patients with disease-related autoimmune, inflammatory and lymphoproliferative complications (Table 1) showed markedly decreased antibody responses compared to those with infection only (2804 vs. 175 BAU/mL median value, P = 0.0003 by Mann-Whitney test, Figure 1H, left panel). This observation, however, held true only for the “early” (<3 months) collection group, as patients from the “late” group (>3 months) did not show differences (Figure 1H, right panel). Furthermore, the correlation between the time elapsed since the administration of the last dose and anti-RBD IgG levels in this sub-cohort remained highly significant, further demonstrating that patients with complications are able to achieve comparable levels of seroconversion through a slower but sustained response (Spearman’s r = 0.63, P < 0.0001, Figure 1I). A cross-sectional overview of all available serology data is presented in Supplementary Figure S2A, where data from 6 patients with X-linked agammaglobulinemia (XLA) and 5 patients with primary unspecific hypogammaglobulinemia (puHG) are included for comparison. As shown in Supplementary Figure S2B–C, anti-RBD IgG titers in XLA and puHG patients were low, with a subset of individuals exhibiting detectable humoral responses that were maintained over time.

### Impact of immunoglobulin replacement therapy on anti-RBD IgG titers

Immunoglobulin (Ig) replacement therapy represents the gold-standard treatment for patients with primary and secondary antibody deficiencies. All patients in the study received intravenous Ig replacement therapy (IVIG), subcutaneous Ig replacement therapy (SCIG), or both (Table 1). Ig products were reported to contain infection-induced antibodies from late 2020 and vaccine-induced antibodies from late 2021 [28], and the RBD-based assay used in this study detects both endogenously produced antibodies and antibodies derived from Ig replacement therapy. This raises the possibility that samples collected at later time points—particularly after the third and fourth vaccine doses—may have been influenced by exogenous Ig, complicating attribution of rising anti-RBD IgG levels solely to endogenous response to sequential vaccine boosting. We therefore examined the impact of exogenous Ig supplementation on the measurement of anti-RBD IgG titers. First, we observed that the contribution of Ig replacement therapy route of administration to anti-RBD IgG detection was limited, as anti-RBD IgG titers did not significantly differ between patients receiving IVIG, SCIG, or a combination of both (Figure 2A). Second, and more importantly, when we analyzed serum anti-RBD IgG levels as a function of the monthly Ig dose administered, we found a strong negative correlation in vaccinated patients without prior SARS-CoV-2 infection (Spearman’s r = −0.44, P = 0.0005; Figure 2B), and no correlation in patients who experienced breakthrough infection prior to sampling (Spearman’s r = −0.10, P = 0.6; Figure 2C). Ig dosing was significantly higher in patients with bronchiectasis, granulomatous–lymphocytic interstitial lung disease (GLILD), and splenomegaly, all conditions associated with lower serum anti-RBD IgG levels (Figure 2D). These observations suggest that exogenous immunoglobulin supplementation was not the primary driver of serum anti-RBD IgG modulation in this study, as patients with weaker vaccine responses received higher Ig doses, yet this supplementation was insufficient to compensate for their impaired endogenous humoral response. Most of the patients for whom Ig replacement data were available experienced SARS-CoV-2 infection between the post-vaccination sampling timepoints. To further discriminate between the effects of Ig supplementation and intercurrent infection, we therefore focused on eight patients who did not experience infection between the two sampling timepoints. Four of these patients had contracted COVID-19 prior to both sampling timepoints and were therefore included in the analysis. As shown in Figure 2E, the change in serum anti-RBD IgG between the two post-vaccination timepoints (differential BAU/mL) did not correlate with the total amount of administered Ig, expressed as grams per kilogram of body weight (g/kg bw), during the same period (Spearman’s r = 0.57, P = 0.15). Together, these data suggest that Ig-replacement therapy did not significantly affect anti-RBD IgG detection and that the endogenous humoral response constitutes the major driver of serum anti-RBD IgG levels in these patients.

**Figure 2.**
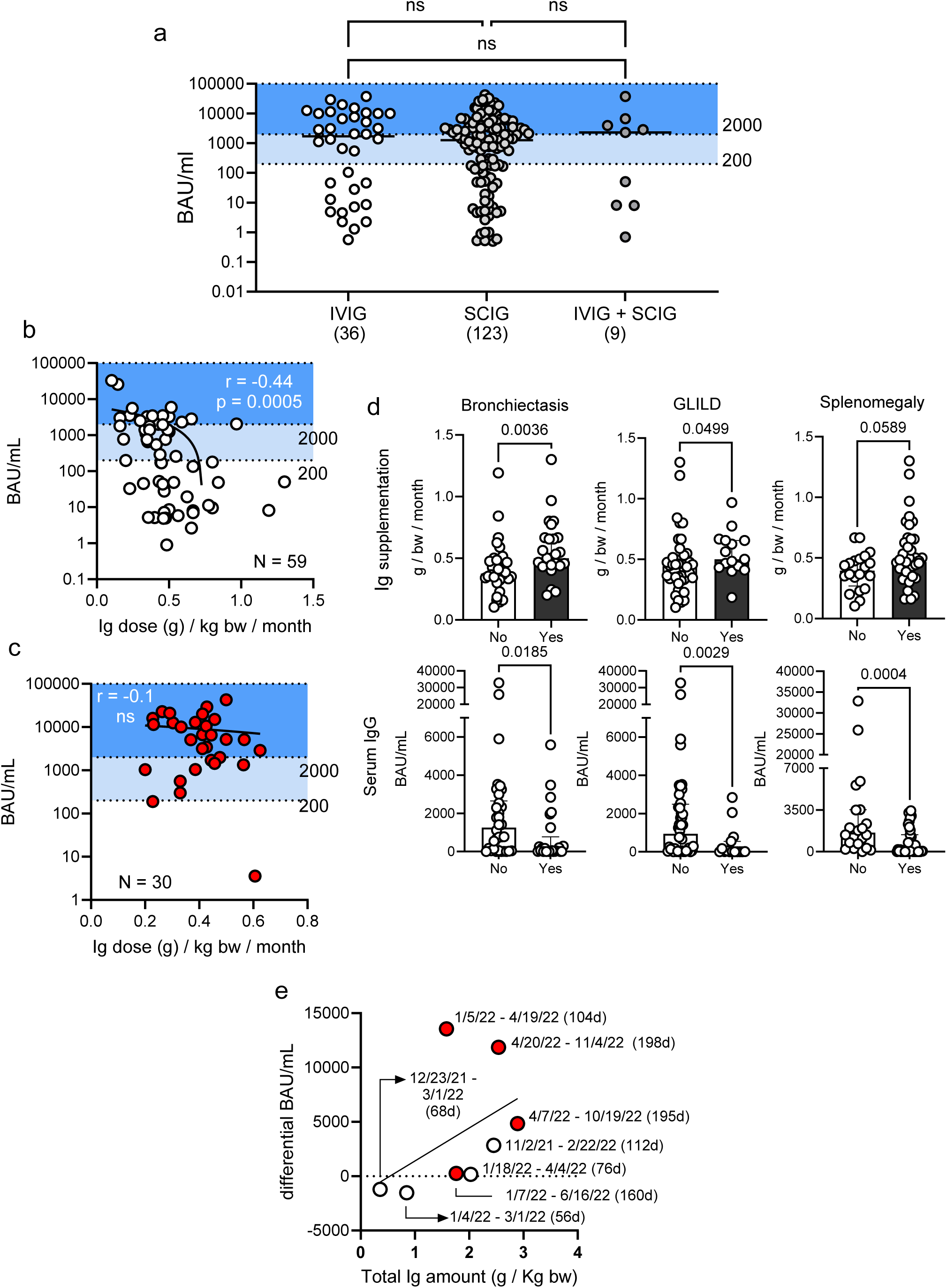
Impact of immunoglobulin replacement therapy on anti-RBD IgG titers. (A) Anti-RBD IgG levels in patients receiving intravenous (IVIG), subcutaneous (SCIG), or combined Ig replacement therapy (ANOVA; not significant). For each patient, measurements from multiple timepoints were included when available. The number of datapoints per group is shown in parentheses. Data from patients with other primary adaptive immunodeficiencies are also included. BAU, binding antibody units. (B–C) Correlation between anti-RBD IgG levels and monthly Ig dose (g/kg body weight/month) in patients without (B) and with (C) prior COVID-19 before sampling. Spearman’s r and P values are indicated in each panel. (D) Comparison of Ig supplementation and serum anti-RBD IgG levels in patients with and without bronchiectasis, granulomatous–lymphocytic interstitial lung disease (GLILD) and splenomegaly. Mann–Whitney test P values are shown. (E) Correlation between the change in serum anti-RBD IgG levels between two sequential sampling timepoints (differential BAU/mL) and the total amount of Ig administered (g/kg body weight) during the same period. Data from patients with infection before sampling are shown in red. Sampling dates and the number of days between samplings are indicated. BAU, binding antibody units; bw, body weight.

### Robust T cell responses upon vaccine exposure in CVID

We next investigated whether variations in serological responses in CVID patients were linked to the dynamics of peripheral B and T cell subsets. After three vaccine doses, anti-RBD IgG levels were positively associated with class-switched memory B cells (Kendall’s τ-b = 0.33; p = 0.020; Supplementary Figure S3A) and inversely associated with circulating follicular helper T cells (Tfh) (Kendall’s τ-b = −0.31; p = 0.020; Supplementary Figure S3B), consistent with early peripheral Tfh homing to secondary lymphoid organs upon boosting. Regulatory T cells (Tregs) showed a positive correlation with anti-RBD IgG titers in these samples (Kendall’s τ-b = 0.29, p = 0.021, Supplementary Figure S3C). CD21low B cells, known as an “inflammatory B cell subtype” [29], did not correlate with humoral response, and no significant association was observed in samples from the late-collection group (data not shown). None of the patients included in these analyses had a history of SARS-CoV-2 infection, confirming that the observed associations reflect vaccine-induced immunity. These results indicate that early B and T cell dynamics are linked to serum IgG levels in CVID.

The observation of a time-dependent coordination between humoral and cellular immunity prompted us to analyze vaccine-induced cell-mediated responses in PBMCs from CVID patients. Using the activation-induced marker (AIM) assay, we observed robust spike-specific responses in both CD4^+^ helper (Th) and CD8^+^ cytotoxic (CTL) T cell subsets following *ex vivo* restimulation with pools of overlapping 15-mer peptides from the spike protein (Figure 3). The median frequency of AIM^+^ Th cells increased from a pre-vaccination baseline (V0) of 0.034% to 0.26% (P < 0.0001, Figure 3A) after two vaccine doses (V2), with responses plateauing after the third dose (V3; 0.17%, P = 0.034; Figure 3A). AIM^+^ CTL frequencies increased from a median of 0.088% at V0 to 0.17% after two doses, although this change did not reach statistical significance (P = 0.074; Figure 3B). Spike-specific polyfunctional T cell responses in CVID patients after two or three mRNA vaccine doses were comparable to those of healthy donors in both the CD4^+^ (0.16% vs. 0.20%, ns; Figure 3C, left panel) and CD8^+^ compartments (0.25% vs. 0.22%, ns; Figure 3C, right panel). Similarly, T cell reactivity remained preserved in samples collected more than 3 months after vaccination (Figure 3D), indicating that cell-mediated immunity in CVID is activated early and is long-lived. Next, we leveraged longitudinal data from six donors who became infected between their last two sampling points to assess post-infection T cell activation dynamics. Representative trajectories from two donors (Supplementary Figure S4A) showed that vaccine-induced T cell activation persisted for several months. As evidenced by changes between the final two sampling points, Th responses plateaued at later time points, whereas CTL reactivity was initially lower but increased progressively and stabilized thereafter (Supplementary Figure S4B). Median frequencies of polyfunctional CTLs also exceeded those of Th cells (0.41% vs 0.14%). Fold-change differences between pre- and post-infection measurements further indicated more sustained reactivity in the CTL compartment compared with the Th compartment (0.5801 vs 3.844; P = 0.0625; Supplementary Figure S4C). Moreover, in samples collected ≥3 months after vaccination, spike-specific CTL frequencies correlated positively with time since the last vaccine dose (Pearson’s R = 0.77; P = 0.0056; Supplementary Figure S4D), with individuals reporting prior SARS-CoV-2 infection exhibiting higher spike-specific CTLs and clustering at later time points. Together, these findings suggest that T cell immunity in CVID patients emerges early after vaccination and remains durable over time.

**Figure 3.**
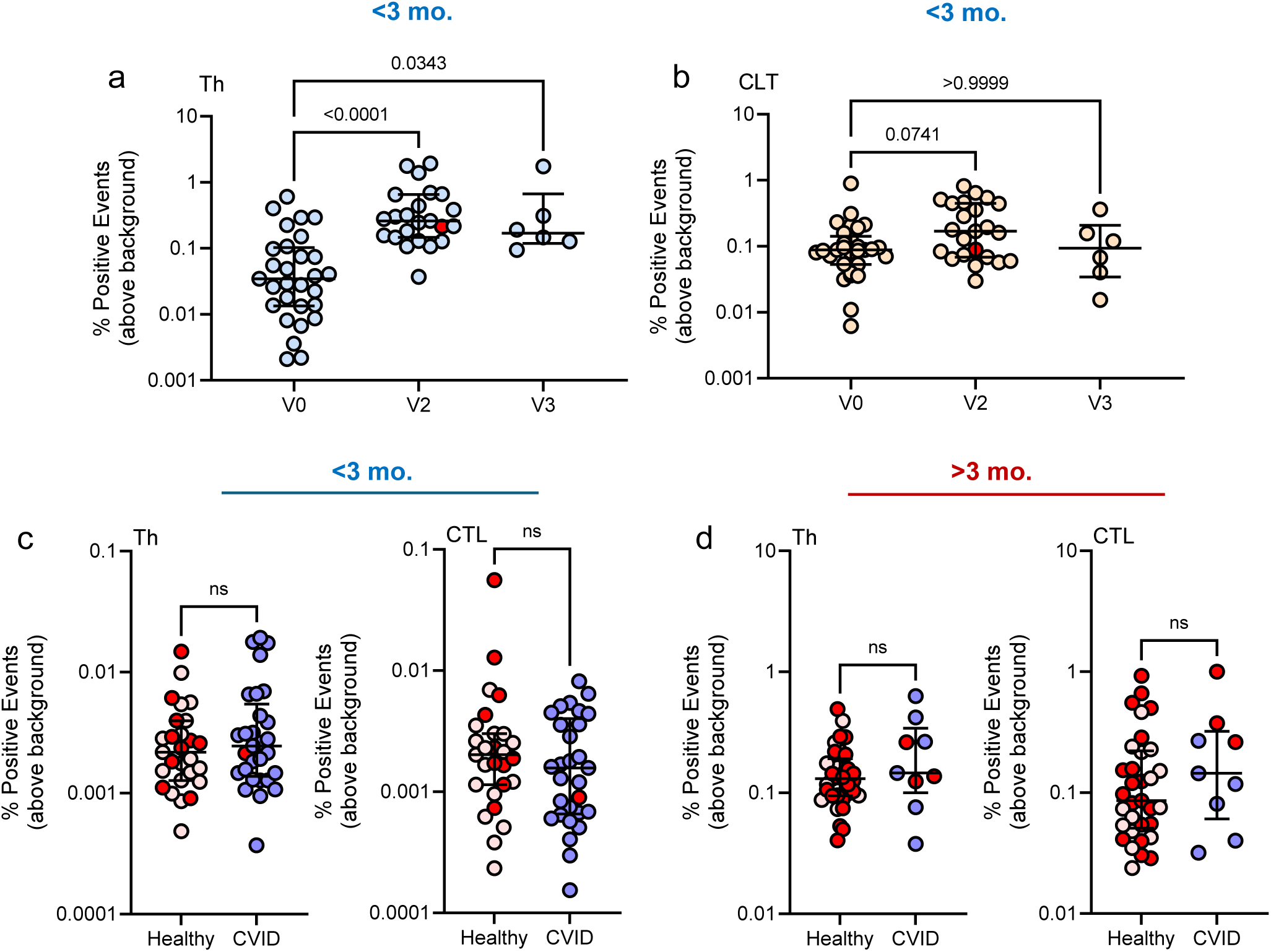
Robust T cell responses upon vaccine exposure in CVID. (A–B) Percentages of activated AIM^+^ polyfunctional CD4^+^ Th (A) and CD8^+^ CTL (B) T cells following stimulation with SARS-CoV-2 PepTivator pools before vaccination (V0) and after the second (V2) or third (V3) dose. All PBMC samples were collected within 3 months post-vaccination. Kruskal–Wallis test; P values shown. (C–D) Percentages of AIM^+^ CD4^+^ Th and CD8^+^ CTL cells following stimulation after two (V2) or three (V3) vaccine doses. Patients with prior SARS-CoV-2 infection are indicated in red. Samples were collected within (C) or after (D) 3 months from vaccine administration. Mann–Whitney test; P values shown. ns, not significant.

## Discussion

In this study, we show that CVID patients mount delayed but boostable humoral responses to SARS-CoV-2 mRNA vaccination, with seroconversion rates increasing progressively with successive booster doses, yet remaining substantially lower than in healthy controls. Dose-dependent IgG increases were detectable only beyond three months post-vaccination, indicating that early serological measurements systematically underestimate vaccine responsiveness in this population. In contrast, spike-specific CD4^+^ and CD8^+^ T cell responses were rapidly induced, durable, and comparable in magnitude to those of healthy donors. Early post-booster IgG levels correlated with class-switched memory B cells and Treg frequencies and inversely with circulating Tfh cells, suggesting active lymphoid tissue engagement during peak vaccine response.

Consistent with previous reports [18, 30–32], early serological responses to two mRNA vaccine doses were markedly reduced in our cohort, with only about one-third of CVID patients achieving seroconversion. Additional doses increased both the proportion of responders and the magnitude of anti-RBD IgG titers, ultimately resulting in seroconversion in most patients. Although this pattern aligns with findings from other studies, our data highlight sampling time as a determinant for detecting serological responses in CVID. In contrast to the rapid seroconversion typically observed in healthy individuals [16], no dose-dependent differences in anti-RBD IgG levels were detectable in CVID samples collected within 3 months of vaccination. A clear, dose-dependent increase emerged only in samples obtained >3 months after vaccination (late-collection group). Anti-RBD IgG titers appeared unstable after two doses but showed a consistent increase after the third dose, an effect that reached significance exclusively in the late-collection group. The positive correlation between IgG titers and time since the last vaccine dose further supports this delayed-response model, indicating that vaccine-induced humoral immunity follows slower kinetics in CVID and stabilizes after the third dose. Together, these findings suggest caution when interpreting serological data obtained from samples collected less than 3 months post-vaccination [18, 33, 34].

Disease-related complications were associated with significantly reduced humoral responses, but this effect was detectable only in the early sampling group. By contrast, sera collected at later time points showed no difference between complicated and uncomplicated CVID, implying that delayed antibody generation may partially compensate even in immunologically more severe phenotypes.

Ig replacement therapy is the standard treatment for individuals with primary antibody deficiencies [35]. These preparations are produced from pooled donor plasma and typically require approximately one year of manufacturing, with finished products having a shelf life of up to three years [36]. Prior to the COVID-19 pandemic, commercial Ig formulations did not contain detectable antibodies against SARS-CoV-2 [37]. However, following widespread infection and vaccination, Ig products manufactured from plasma collected after early 2021 began to contain substantial levels of anti–SARS-CoV-2 antibodies [28, 38, 39].

The assay used in this study detects both endogenous and exogenous antibodies, raising the possibility that passive antibody transfer might contribute to measured anti-RBD IgG levels. However, several observations argue against this interpretation. First, anti-RBD IgG levels did not differ according to the route of Ig administration (IVIG versus SCIG), which have large differences in Ig kinetics after administration [40]. Second, anti-RBD IgG levels showed a negative correlation with administered Ig dose in infection-naïve patients, inconsistent with Ig replacement therapy being the dominant source of the measured antibody signal, and instead likely reflects clinical dose adjustment whereby patients with more severe immune dysfunction receive higher Ig doses but mount weaker endogenous vaccine responses, or a suppressive effect of high-dose Ig administration on B cell function [41]. Furthermore, longitudinal analyses in patients without intercurrent infection showed no relationship between cumulative Ig exposure and changes in anti-RBD IgG levels over time. Together, these findings indicate that the serological responses observed in this cohort primarily reflect endogenous immune responses rather than passive antibody transfer from Ig replacement therapy.

T cell responses are central to antiviral defence, particularly when antibody availability is limited [42–44]. Tfh typically show increased frequencies in blood alongside higher PD-1 expression in CVID, often associated with a Th1 profile and, at least in some cases, impaired B cell maturation [45]. Phenotypic analyses of peripheral B and T cell subsets revealed immune correlates that aligned with the temporal shift in humoral responsiveness. Early after vaccination, serological activity aligned with lower frequencies of circulating Tfh cells and higher levels of class-switched memory B cells, suggesting an active role of the CD4^+^ compartment in vaccine responsiveness [23, 34]. This interpretation was further supported by the inverse correlation observed between circulating Tfh cells and antibody levels in early samples, potentially reflecting the homing of recirculating Tfh pools to lymphoid organs after vaccine rechallenge. Notably, these associations were time-dependent (early- vs. late-collection groups), reinforcing the idea that immune-system functionality should be interpreted in the context of sampling timing.

When we evaluated vaccine-specific Th-cell responses using spike-peptide stimulation and AIM assay, we found that T cell immunity elicited by mRNA vaccination in CVID patients emerged early and was durable. This aligns with several studies reporting high rates of polyfunctional activation in nearly all CVID patients 4–6 weeks after dose 2 [23, 34]. Other studies, however, have described defects in Th responses to spike peptide stimulation and either failed to detect CTL activation or reported weak CTL responses [32, 46–48]. In our cohort, activation increased in both CD4^+^ and CD8^+^ compartments following vaccination and remained detectable for several months, with magnitudes comparable to those of healthy individuals. Differences across studies may reflect differences in assay sensitivity (e.g., FluoroSPOT, tetramers), the use of 15-mer peptide pools, which are known to favor Th responses [49], or enhanced detection afforded by the polyfunctional AIM analysis used in this study. Furthermore, in six donors who became infected between their last two sampling timepoints, cytotoxic T cell responses not only persisted but, in some cases, modestly increased following SARS-CoV-2 infection. These results are in line with previous reports showing that in healthy individuals vaccination primarily induces rapid and robust CD4^+^ T cell responses, whereas CD8^+^ T cell responses develop more gradually and are more prominently expanded following infection or repeated antigen exposure [50]. The positive association between presence of activated CTLs and time since vaccination suggests the preservation of a sustained cell-mediated immune program, even when humoral responses are weak or delayed. Consistent with findings from other cohorts of immunocompromised donors [44], these observations highlight that T cell immunity can help compensate for suboptimal antibody production in these patients and likely contributes to protection.

This study has limitations. Some sub-analyses included a limited number of patients and should therefore be interpreted with caution. In addition, functional data on the capacity of purified B cells from CVID patients to mount IgG responses to the tested vaccines are lacking.

## Conclusions

Taken together, these findings highlight that immune responses in CVID should be interpreted in the context of delayed humoral kinetics and preserved cellular immunity. The observation that antibody responses in these patients may only become detectable at later timepoints after vaccination suggests that early serological measurements can underestimate vaccine responsiveness in this heterogeneous patient population. At the same time, the presence of robust and durable antigen-specific CD4^+^ and CD8^+^ T cell responses indicates that cellular immunity may provide an important layer of protection when antibody production is impaired. More broadly, these findings support the continued use of booster vaccination strategies in patients with primary antibody deficiencies and emphasize the importance of integrating cellular immune readouts and appropriate sampling intervals when evaluating vaccine responses in immunocompromised populations.

## Statements and Declarations

### Author Contributions

LF, RØL, BF, KL, PA, and LAM conceived and designed the study. LF, QKL, JRO, VC, KPL, LO, and FLJ performed the experiments. LF drafted the first version of the manuscript. LF, BF, LAM, and RØL revised and finalized the manuscript. BF, RØL, IN, TS, SFJ, MSAF, and PA contributed to patient recruitment, clinical characterization, and sample collection. All authors contributed to data analysis, critically reviewed the manuscript, and approved the final version.

### Data Availability

The datasets generated and analyzed during the current study are available from the corresponding author upon reasonable request. Individual-level clinical data are not publicly available due to privacy regulations and ethical restrictions related to patient confidentiality.

### Ethics Approval

This study was performed in line with the principles of the Declaration of Helsinki. The study protocol and all associated biobanking procedures conducted at Oslo University Hospital were approved by the Norwegian Regional Committee for Medical and Health Research Ethics (approval number 233704) and by the Data Protection Officer of the Oslo University Hospital.

### Consent to Participate

Written informed consent was obtained from all individual participants included in the study.

### Consent to Publish

The authors affirm that all participants provided written informed consent for publication of data included in this manuscript.

## Acknowledgements

We thank the patients and healthy donors who participated in this study. We are grateful to the clinical and technical staff at the Department of Immunology, Oslo University Hospital, for assistance with sample collection and processing. We also thank members of the research groups involved for helpful discussions and technical support.

