## Supplementary Figures for "Delayed humoral kinetics but stabilization of IgG responses in common variable immunodeficiency after SARS-CoV-2 mRNA booster vaccination"

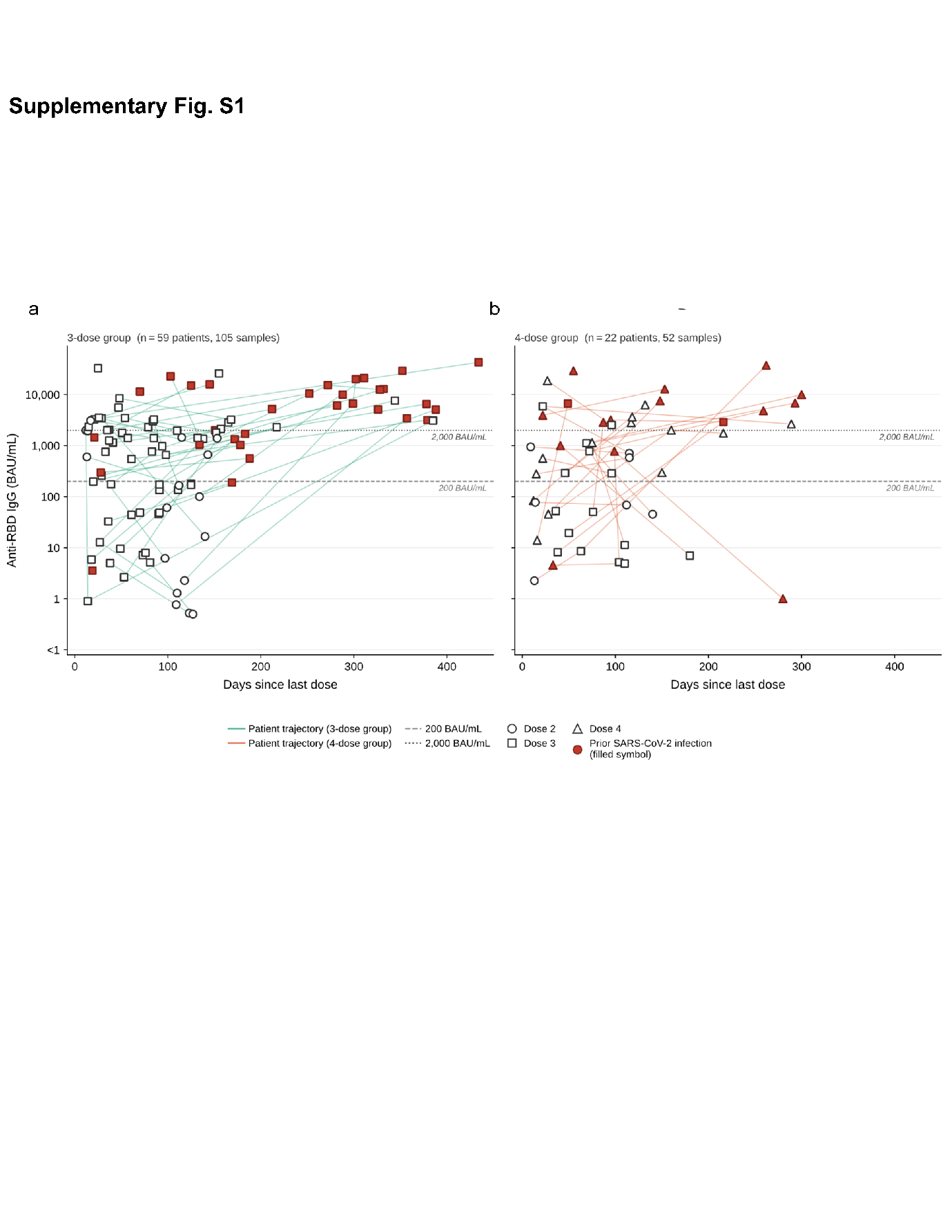

**Supplementary Fig. S1. Longitudinal anti-RBD IgG responses according to vaccine dose and time since vaccination** (**A–B**) Serum anti-RBD IgG levels (BAU/mL) in patients receiving up to three (A) or four (B) vaccine doses. In (A), a subgroup of patients (n = 44/59) was sampled only after the third dose, as they were enrolled after receiving two doses. The number of vaccine doses received prior to sampling is indicated by the symbols shown below the figure. The x-axis represents days since 25 May 2021 (day 0). Patients with prior SARS-CoV-2 infection before vaccination are shown in red.

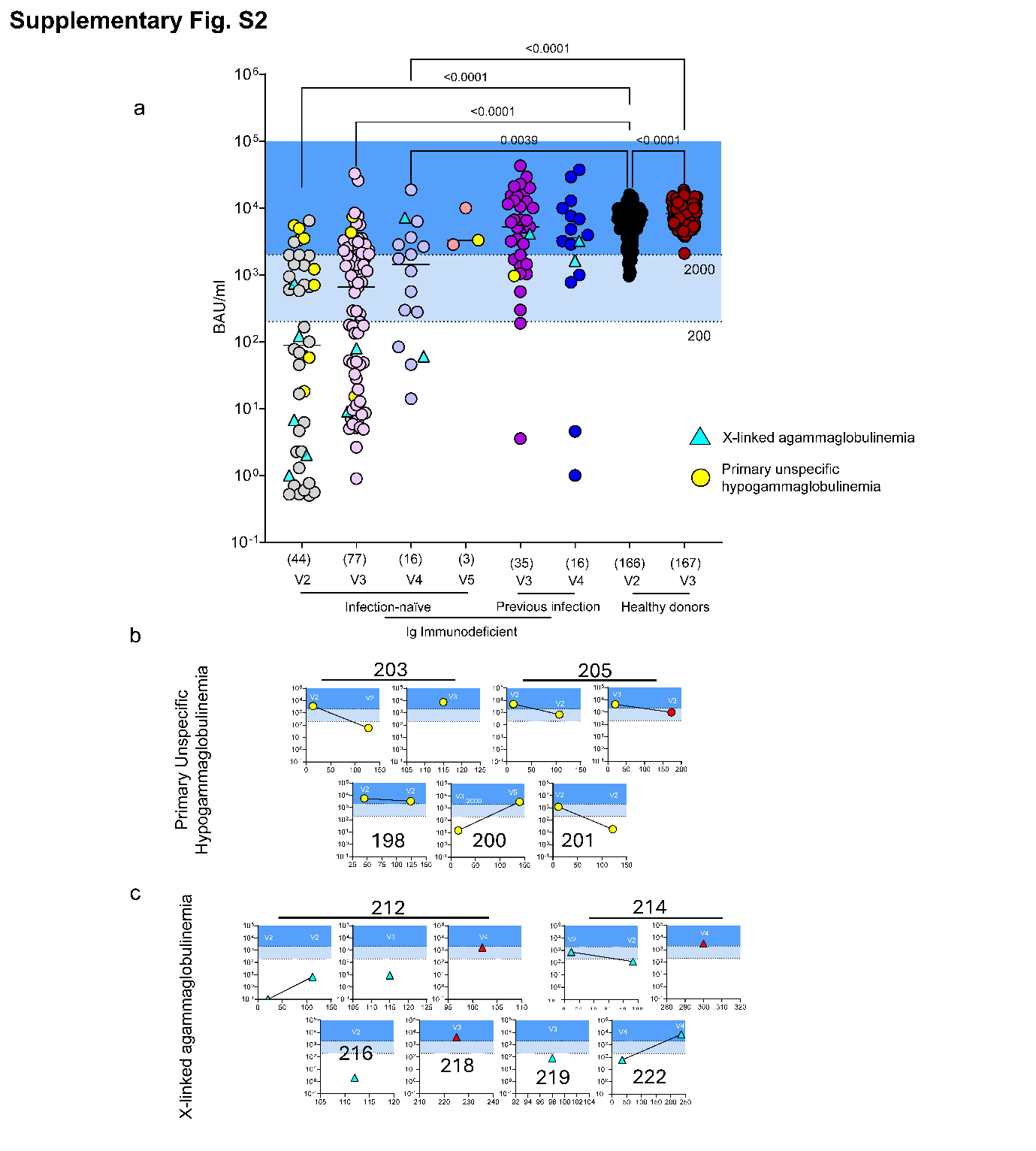

**Supplementary Fig. S2. Cross-sectional overview of serological data (A)** Seroconversion rates in healthy donors and in patients with CVID, X-linked agammaglobulinemia (XLA), or other primary adaptive immunodeficiencies, with and without prior SARS-CoV-2 infection and across different mRNA vaccine doses. Blue-shaded areas denote seroconverted individuals (>200 BAU/mL) and strong responders (>2000 BAU/mL). Kruskal–Wallis test; P values are shown. Numbers of patients in each group appear in parentheses along the X-axis. **(B–C)** anti-RBD IgG levels in other primary adaptive immunodeficiencies (B) or in patients with XLA (C). Vaccination status and days elapsed since the last dose are shown for each patient. Red symbols indicate post-infection values. BAU: binding antibody units.

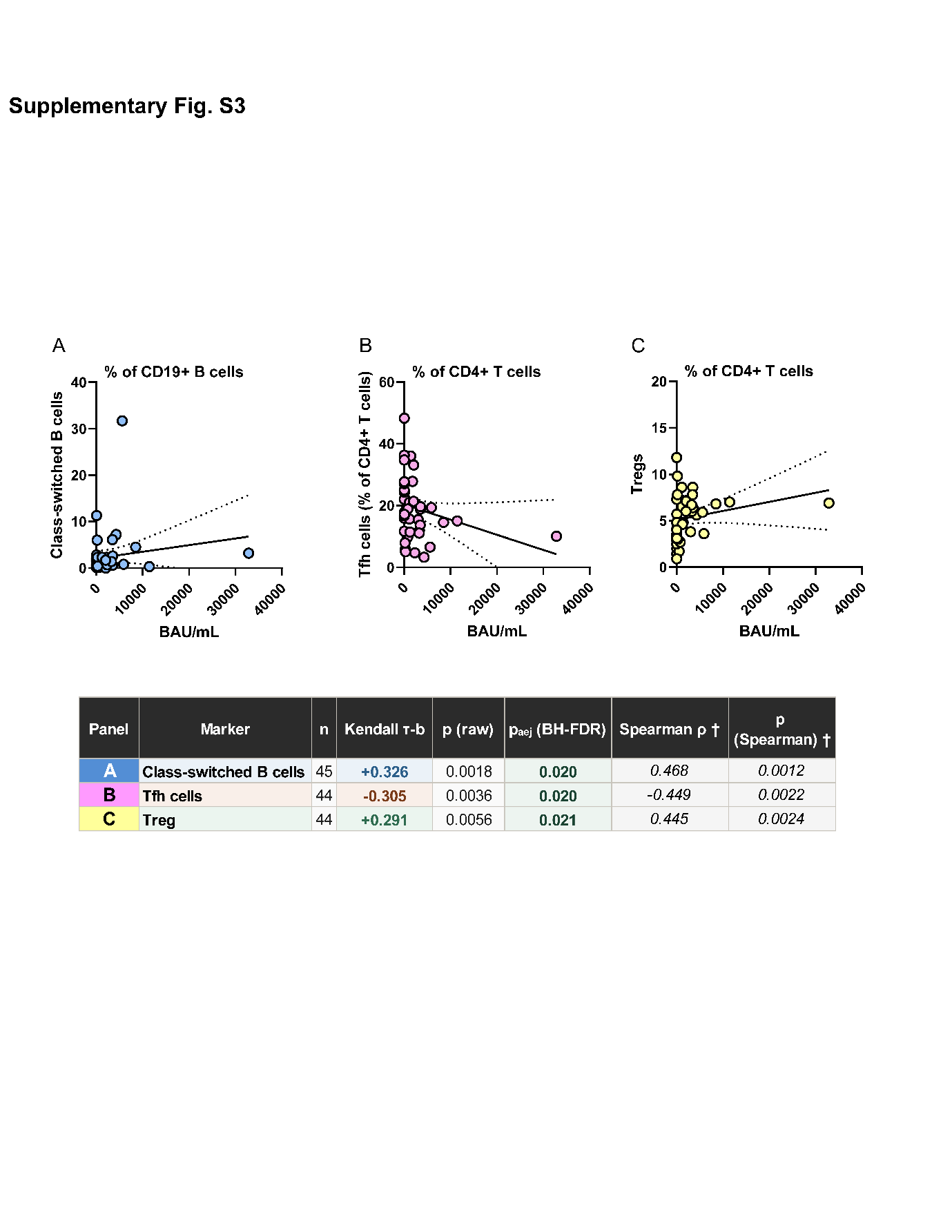

**Supplementary Fig. S3. Peripheral dynamics of T and B cell subsets correlate with the magnitude of humoral response** Scatter plots of anti-RBD IgG serum concentration against immunophenotyping markers in CVID patients sampled ≤90 days after a third vaccine dose (early post-booster group). **(A)** Class-switched B cells (% of CD19⁺ B cells), **(B)** Tfh cells (% of CD4⁺ T cells), **(C)** Treg (% of CD4⁺ T cells). The x-axis displays anti-RBD IgG (BAU/ml) on a log₁₀ scale; the y-axis shows the immunophenotyping marker as a percentage of the parent gate (linear scale). Each circle represents one donor (n = 44–45 per panel after pairwise exclusion of missing values). The results of 3 out of 11 correlations tested across four subgroups (44 tests total) that survived Benjamini–Hochberg FDR correction at the 5% level are shown.

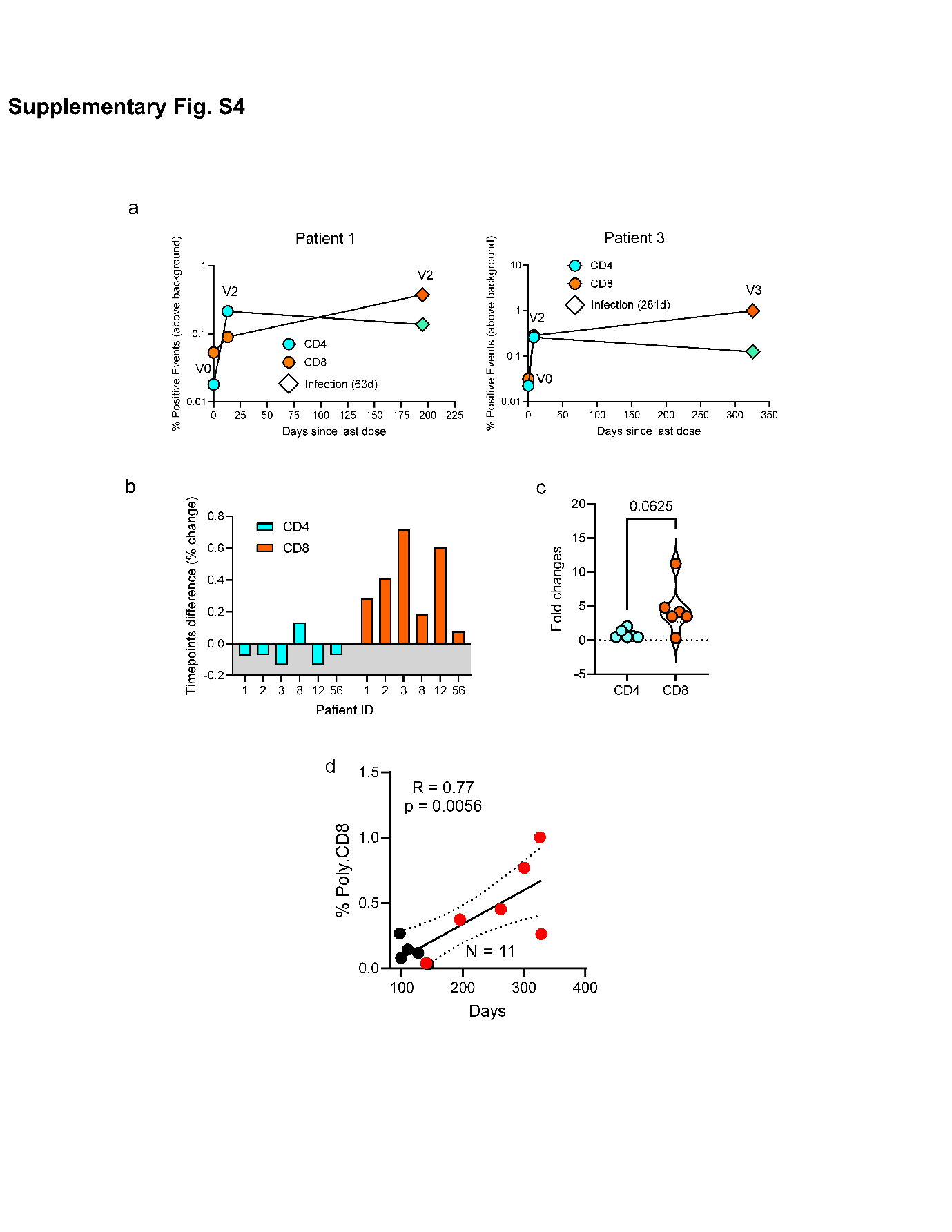

**Supplementary Fig. S4. Sustained CTL response in CVID (A)** Activated AIM⁺ Th and CTL responses plotted over time since the last vaccine dose. Squared symbols denote post-infection values (days post-infection indicated). Two representative patients (1060, 1065) are shown. **(B)** Differences in percentages of AIM⁺ polyfunctional Th and CTL cells between the last and previous timepoint. Gray shading denotes negative differences. **(C)** Fold changes in AIM⁺ cell frequency between the last and previous timepoint for each patient (N = 6). **(D)** Correlation between activated AIM⁺ CTLs and time since the last vaccine dose in patients sampled ≥3 months post-vaccination (N = 11). Red-circled dots indicate post-infection values. Pearson’s R and P values are shown. Wilcoxon test; ns = not significant. All percentages are background corrected.
